# Novel markers to detect HER2 amplification in Breast cancer

**DOI:** 10.1101/2021.11.20.21266647

**Authors:** Shikha Mudgal, Arnav Kalra, Bina Ravi, Shalinee Rao, Nilotpal Chowdhury

## Abstract

Overexpression of HER2 in breast cancer is an important prognostic and predictive biomarker, assessed using immunohistochemistry (IHC) and in situ hybridization (ISH). More than 20% of tumours are graded equivocal on IHC and is send for reflex testing via ISH. In situ hybridization (ISH) is an expensive assay and is not available widely in resource limiting areas. Therefore, we propose that genes found significantly co-expressed with HER2 in breast cancer can be used as surrogate markers for HER2 in breast cancer which can detect HER2 positivity on IHC itself. This hypothesis is based on analysis of publicly available datasets from the Gene Expression Omnibus (GEO) database. The genes found most significantly correlated with HER2 expression were PGAP3 (r = 0.85), GRB7 (r = 0.82), STARD3 (r = 0.78), CDK12 (r= 0.68), PSMD3 (r =0.67) and GSDMB (r = 0.63). We hypothesize that these identified surrogate markers for HER2 amplification which can be detected on IHC can detect HER2 amplification status in HER2 equivocal tumors based on IHC staining alone and will reduce the number of HER2 2+ (equivocal) category tumours.

## INTRODUCTION

Human epidermal growth factor receptor-2 (HER2) is amplified in around 20 to 25% of breast cancers.[1]. This amplification is associated with an aggressive phenotype with poor prognosis,but which is also amenable to treatment by anti-HER2 treatment [2]. HER2 assessment is a key to personalized treatment, especially with HER2 targeted antibody trastuzumab or pertuzumab, [3] but the treatment options are limited by high financial load.[4] Hence, accurate estimation of IHC status is essential.

The recommended protocol for estimation of HER2 status in breast cancer includes Immunohistochemistry (IHC) as an initial step, followed up, if needed by Fluorescent (or Chromogenic) in-situ hybridization (ISH). Tumour blocks are usually tested first by IHC and graded as 0, +1, +2 and +3. Scores 0 and +1 are considered negative whereas + 3 is considered positive. Tumors having IHC 2+ scores are considered tumors of equivocal HER2 amplification status, and which have to be tested again by single or dual probe in situ hybridization (ISH) assay for assessment of the HER2 status [5]. While ISH are considered the “Gold-standard”, they are costly and not available widely in many resource poor settings due to the high costs and lack of technical expertise. In this setting, it makes sense economically to examine other surrogate markers of HER2 amplification which have potential to reduce the number of equivocal cases on IHC, which are cheaper and require lesser technical expertise. Here, we suggest a set of novel IHC markers which may be used to predict the HER2 status of breast cancer. We start with a description of the method analysis conducted to come to our hypothesis, followed by the results, which will lead to the hypothesis and implications.

## METHODS

NCBI-GEO (Gene Expression Omnibus) database was searched for HER2 positive breast cancer gene expression series in *<u>Homo sapiens</u>*. We found three datasets for breast cancer patients who had not undergone any systematic therapy,(GSE11121 [6], GSE2034 [7] and GSE7390 [8]). The Clinical characteristics of the patients in the data series are given in Table 1. The raw data (.CEL files)of these three datasets were downloaded from NCBI-GEO database. These file were then normalized using frozen robust multi-array average (fRMA) [9] Bioconductor package in R version 3.6.6. Probes of the normalized files were collapsed to gene symbols using WGCNA Bioconductor package. Pearson correlation coefficient for each gene in the matrix of all the data sets GSE11121, GSE2034 and GSE7390 were calculated with HER2 gene (ERBB2), using in the statistical environment R v 3.6.6.

**Table 1.** Summary of patient’s characteristics of the three series analysed.

|  |  | GSE2034(N=286) | GSE11121(N=200) | GSE7390(N=198) |
| --- | --- | --- | --- | --- |
| Median follow up period in years.<br>(range) |  | 8.8 (0.17 – 14.25) | 8.6 (0.08 – 20.00) | 13.7(0.34 – 24.95) |
| Mean patient age in years (range) |  | 54 (26 – 83) | 60 (25 - 90) | 46 (24 – 60) |
| Radiotherapy given | Yes | 248 | 125 | - |
|  | No | 38 | 75 | - |
|  | unknown | 00 | 00 | 198 |

## RESULTS

Top most correlated genes with a correlation coefficient above 0.6 were extracted from the combined dataset (Table 2). These genes were PGAP3,GRB7, STARD3, CDK12, PSMD3 and GSDMB All these genes were found to be localized to an area near the locus of HER2 in chromosome 17.

**Table 2:** The location of the top genes correlated with HER2.

| Genes | Correlation<br>coefficient | Protein | Subcellular location | Chromosome location on<br>GRCh38 reference genome |
| --- | --- | --- | --- | --- |
| ERBB2 | - | HER2 | Cell membrane | 17q12<br>(39688084..39728662) |
| PGAP3 | 0.85 | Post-GPI attachment to<br>proteins factor 3 | Endoplasmic reticulum<br>membrane, Golgi apparatus<br>membrane | 17q12<br>(39671122..39688070) |
| GRB7 | 0.82 | Growth factor receptor-<br>bound protein 7 | Plasma membrane | 17q12<br>(39737938..39747285) |
| STARD3 | 0.78 | StAR-related lipid<br>transfer protein 3 | Late endosome membrane | 17q12<br>(39637080..39664201) |
| CDK12 | 0.68 | Cyclin dependent<br>kinase 12 | Nucleus<br>Nucleus speckle | 17q12<br>(39461486..39567560) |
| PSMD3 | 0.67 | 26S Proteasome non-ATPase regulatory<br>subunit 3 | Cytosol, nucleosome,<br>nucleus | 17q21.1 |
| GSDMB | 0.63 | Gasdermin - B | Cytosol, Cell membrane | (39980807...39997959) |

The scatterplot of the expression of the three top genes with HER2 is given in Figure 1.

**Figure 1:**
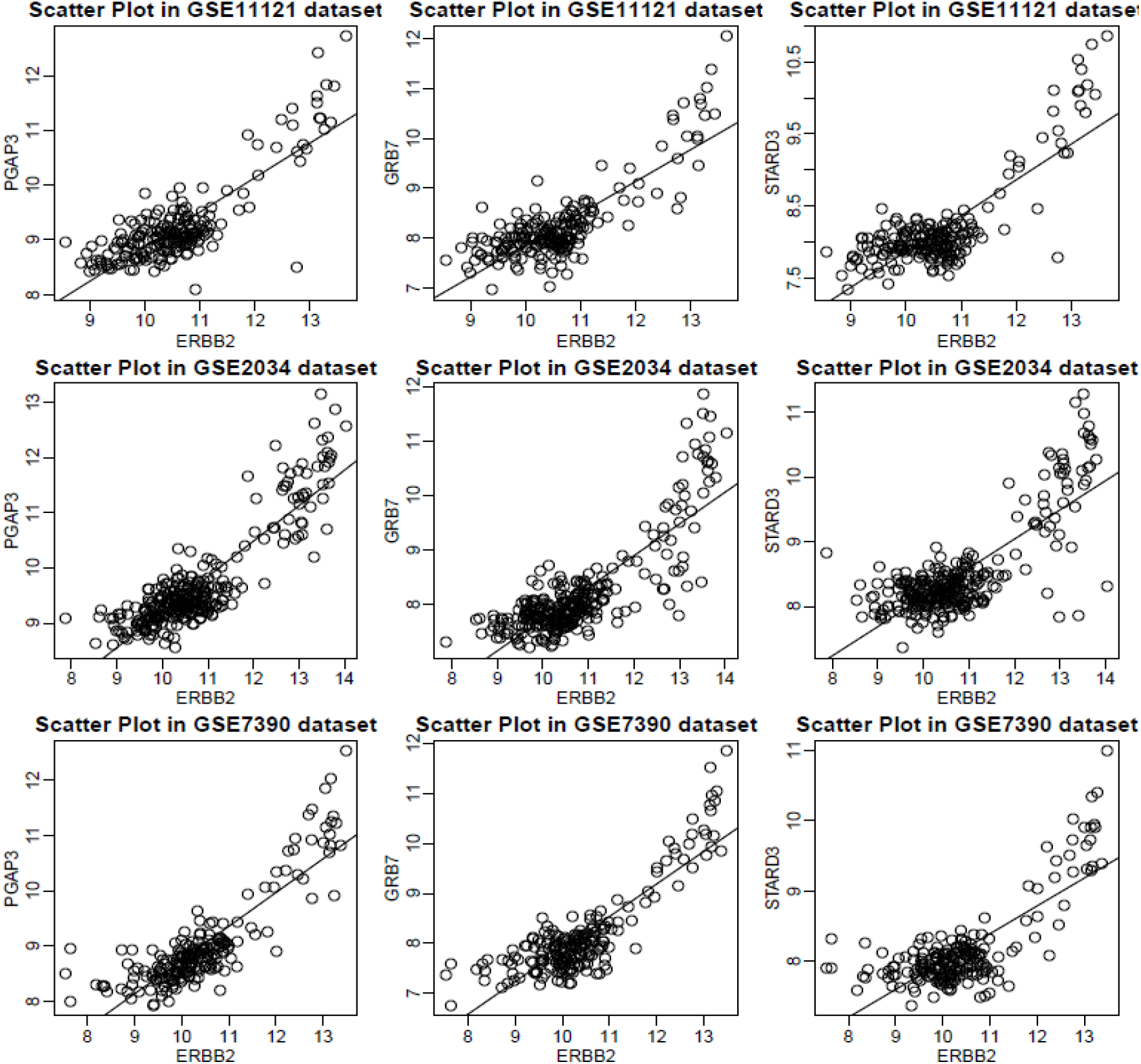
Scatter plot showing correlation of PGAP3, GRB7 and STARD3 genes with ERBB2 genes in GSE11121, GSE2034 and GSE7390 datasets.

## DISCUSSION

We hypothesise that IHC of the obtained HER2 co-amplified proteins (including PGAP3, GRB7, STARD3, CDK12, PSMD3 and GSDMB) will result in better discrimination of HER2 amplified breast cancer patients from HER2 non-amplified breast cancer patients. Breast cancer sections showing high expression of these markers with strong intensity signal should discriminate between HER2 amplified and HER2 non-amplified cancers in tumors having equivocal HER2 immunostaining. Increased strong intensity signals of two or more such markers should result in greater specificity in detecting HER2 amplified tumors in such equivocal cases. Therefore, IHC of these HER2 co-amplified proteins on breast cancer should result in reduced number of cases needing ISH due to the better discriminatory ability. Since the cost of IHC is significantly lesser than ISH both in terms of reagents as well as equipments and manpower, this may result in greater cost-effectiveness of HER2 assessment. This should enable significant cost-savings in resource-limited settings.

The reasoning behind the above hypotheses is that the genes adjacent to HER2 on chromosome 17 also get amplified in HER2 amplified tumors. Genomic studies have characterized such “amplicons”. Besides predicting HER2 status, these genes may themselves be play an important role in the natural history of breast cancer.

The hypothesis can be confirmed by doing a diagnostic validation study comparing the IHC biomarkers of the gene products of the HER2 co-expressed genes with HER2. A significant subset of this validation study should be consist of HER2 equivocal (2+) cases. The equivocal cases should be confirmed by in-situ hybridization. If the expression of the IHC positivity of the top HER2 co-expressed genes significantly predicts the HER2 status in IHC equivocal cases of HER2, and if found to be significantly correlated with the IHC staining pattern of HER2, the hypothesis will be deemed to be supported by evidence to put in regular diagnostic use. A further implication of the hypothesis will be that a reference probe targeting these proteins should also result in a good alternative chromosome 17 reference probe for cases found equivocal even after In-situ hybridization for HER2.

## Data Availability

All data produced in the present study are available upon reasonable request to the authors

